# Stay Home and Stay Active? The impact of stay-at-home restrictions on physical activity routines in the UK during the COVID-19 pandemic

**DOI:** 10.1101/2021.01.31.21250863

**Authors:** Victoria Eshelby, Muhammed Sogut, Kate Jolly, Ivo Vlaev, Mark T. Elliott

**Affiliations:** Institute of Digital Healthcare, WMG, University of Warwick, Coventry, CV4 7AL, UK; Institute of Applied Health Research, University of Birmingham, Birmingham, B15 2TT, UK; Warwick Business School, University of Warwick, Coventry, CV4 7AL, UK

**Keywords:** COVID-19, Physical Activity, Step-count, Restrictions, Lockdown

## Abstract

Government restrictions applied during the COVID-19 pandemic in the UK led to the disruption of many people’s physical activity routines, with sports and leisure facilities closed and outdoor exercise only permitted once per day. In this study we investigated which population groups were impacted most in terms of reduced physical activity levels during these periods, and which groups benefitted in terms of increasing their usual level of physical activity. We surveyed UK residents, sampled through users of a rewards-for-exercise app (Sweatcoin; n=749) and an online panel (Prolific; n=907). Of the app users, n=487 further provided daily step-count data collected by the app, prior to, and during the periods of restrictions between March and June 2020. Regression models were applied to investigate factors associated with subjective change (perceived change in physical activity) and objective change (log-percentage change in daily step-count) in physical activity during the periods of restrictions. ANOVAs were used to further investigate the significant factors identified. Key factors associated with a substantial subjective reduction in physical activity included those classed as obese, gym users and people living in urban areas. All participants had a reduced step count during restrictions, with Black, Asian and minority ethnic (BAME) groups, students and urban dwellers showing the largest reductions. Therefore, targeted interventions are required to ensure that the physical and mental health impacts of sedentary behaviour are not exacerbated over the long-term by significant reductions in physical activity identified in these groups, particularly those who are also more vulnerable to the COVID-19 virus.

## INTRODUCTION

Throughout the period of the COVID-19 pandemic, the UK government introduced restrictions as a means to slow the progression of the outbreak. The first phase of restrictions was applied from 23 March 2020 with a ‘Stay at Home’ message. Travel was limited to all but essential journeys, and all non-essential services, including sports and leisure facilities were closed; outdoor exercise was permitted once per day [1]. On the next phase of lockdown restrictions (introduced May 13^th^, 2020), the government re-opened outdoor public places, allowed people to exercise more than once a day and to drive to outdoor destinations. However, gyms and sports facilities remained closed [2].

These unprecedented restrictions led to substantial disruption to many people’s physical activity (PA) routines. Those who relied on gyms and sports facilities were required to find alternative forms of PA that complied with the restrictions. Overall, the disruption is likely to have led to reduced PA and exercise routines, which could potentially increase susceptibility to the COVID-19 as well as increase the likelihood of underlying conditions [3]. Thus, it is essential to identify populations that are most impacted from the restrictions on their PA routines such that future interventions can be developed to help them adapt and maintain or increase PA levels.

In contrast, some people have gained new opportunities in terms of PA during the periods of lockdown. A large proportion of the population switched to working from home [4], while many others were furloughed [5]. The switch in working patterns, for some, increased time for PA, that wasn’t possible previously, for example through the time saved by not commuting to the office.

In this study, we investigate in detail which groups have been positively and negatively impacted in terms of their PA routines due to the UK government restrictions put in place between March and June 2020. Importantly, we have used both self-reported and objective measures of activity change to identify the factors associated with PA change. Furthermore, we identify how the types of PA have changed during the two phases of restrictions, and whether respondents were likely to stick with new or old routines once restrictions were lifted.

## METHODS

### Participants and Timing

Participants were recruited via a physical activity incentives app (Sweatcoin [6]; n=1322) and a survey panel (Prolific [7]; n=932). After removal of duplicate and incomplete entries, 1656 survey responses were used for analysis of subjective measures (app: n=749, panel: n=907). All participants resided in the UK.

Sweatcoin users were further given the option of providing their historic step count data recorded by the app, between 1^st^ February 2020 and the date of completing the survey; 950 users consented to providing this data in addition to their survey responses. After matching data to survey responses and removing entries with more than 50% of days with missing step-count values, 487 participants were used for analysis of objective data (in combination with their survey responses).

The survey ran May 29^th^ to June 10^th^ 2020.

### Ethics

The study was given ethical approval by the Humanities and Social Sciences Research Ethics Committee at the University of Warwick. Informed consent was given by participants before proceeding with the survey questions. Each participant was provided with a nominal payment of £2 for fully completing the survey, which took approx. 10-15 minutes. Participants provided additional consent for sharing step-count data, by ticking a box on the consent form.

### Questionnaire Design

The questionnaire consisted of demographic, wellbeing, physical activity, working status, covid-19 status and opinions, and personality information. The following variables were collected in the survey data (A full sample breakdown is given in Supplementary Information A).

#### General Demographics

We captured gender, age, height, weight and ethnicity information. In addition, we gathered participants’ geographic location using the initial part of their postcode, along with details on whether they lived in an urban, suburban or rural location and whether they had access to a private garden. Finally, we captured whether they had children (under the age of 18) living at home.

#### Wellbeing

We used the four measures of personal wellbeing (Office of National Statistics, [8]) to measure subjective measures of Life Satisfaction, Worthwhile, Happiness and Anxiety on a scale of 0 (not at all) to 10 (completely). In addition, we asked users to rate their overall health on that day, on a scale of 0 (worst health) to 100 (best health).

#### Working status

We asked participants about their current work status over the past week, in terms of whether they were working from home, working at their usual location (away from home), furloughed or not in employment.

#### COVID-19 status and opinions

We asked participants how worried they were about coronavirus and to rate their certainty on whether they had or previously had Covid-19 (or not). Participants further stated whether they had a received a letter stating that they should follow shielding guidelines, and whether they were complying with this. Similarly, we captured the proportion of people participants thought were complying with social distancing measures and government-imposed restrictions of movements.

#### Personality

The Big-Five personality dimensions (openness to experience, conscientiousness, extraversion, agreeableness, neuroticism) [9] were captured using the Ten Item Personality Measure (TIPI; [10]).

#### Physical Activity

Participants were asked the following regarding their physical activity routine:

##### Time spent on activities

Number of hours spent weekly on physical exercise (e.g. swimming, jogging, football, aerobics, gym), cycling and walking.

##### Types of exercise

We asked participants to define the main form of exercise they routinely participated in over the three time periods.

##### Likelihood to stick with new routine

Participants rated how likely they were to return to their original physical activity routine (prior to restrictions) or their new routine (during the second phase of restrictions) once all restrictions were lifted and business had reopened.

##### Employment related physical activity

Participants reported the level of activity they did in work, prior to the restrictions, based on their job role (as defined by the General Practice Physical Activity Questionnaire, GPPAQ [11]).

##### Commuting related physical activity

Participants were asked to state the number of minutes spent walking, cycling, using public transport and driving, during their commute to work.

##### Perceived change in physical activity

Finally, participants were asked to consider their PA based on the three periods relating to times prior to lockdown and two periods of UK government restrictions between March and June 2020 (Table 1). Participants were asked to think about their typical routine, based on the survey period the question referred to. The primary dependent variable we used for analysis was based on perceived change in PA during Phase 1 and Phase 2, relative to the Pre-restriction period. This was based on a Likert scale of −5 (substantially reduced) through to +5 (substantially increased).

**Table 1.** Based on the level of restrictions, three periods were analysed. The date range is based on the start/end date of restrictions coming into effect, based on UK Government announcements [1,2]. Survey period is the time period we asked participants to consider when making their responses. Step count period is the time period over which daily step-counts were analysed for that period.

| Period label | Date range | Survey period | Step count period |
| --- | --- | --- | --- |
| Pre-restrictions<br>(Baseline) | Prior to 23<sup>rd</sup> March 2020 | Week before 16<sup>th</sup> March 2020 | February 1<sup>st</sup> to February 29<sup>th</sup> 2020 |
| Lockdown (Phase 1) | 23<sup>rd</sup> March to 12<sup>th</sup> May 2020 | A typical week between 23<sup>rd</sup> March – 12<sup>th</sup> May 2020 | 23<sup>rd</sup> March to 12<sup>th</sup> May 2020 |
| Relaxed Restrictions<br>(Phase 2) | 13<sup>th</sup> May 2020 onwards | Week prior to survey completion date (29<sup>th</sup> May to 10<sup>th</sup> June 2020) | 13<sup>th</sup> May 2020 to survey completion date. |

### Step Count Data

Historic step count data recorded by the Sweatcoin app [6], was provided by a subset of participants between 1^st^ February 2020 and the date of survey completion. Step count data with more than 50% of days with missing step-count values, were removed. The data was split into three time periods, similar to the survey (see Table 1).

Within each period, daily step count data was averaged across days of the week, resulting in seven mean daily step-count values (Sunday-Saturday) for each participant, per period. To measure proportional change in step-count during the phase 1 period of restrictions, we divided the mean daily step counts in phase 1 by the corresponding value in the baseline period. The natural log of the resulting values was calculated and then the mean taken to get a final phase 1, log-percentage change for each participant. The same procedure was applied to the phase 2 data to get a corresponding value for this period.

## ANALYSES

All analyses were completed using the R programming language (v3.6.2; [12]). Multiple regression models were used to analyse the factors associated with PA change during the lockdown period. We investigated both subjective (perceived change in PA) and objective (log-percentage change in mean daily step count) measures of change as dependant variables, with the survey data used as predictor variables. Continuous variables were standardised, by mean-centering and scaling by the standard deviation. Significant variables were defined as p<0.05. Multicollinearity was tested for between variables using the variance inflation factors (VIF) method; we report the maximum value (VIF_max_) from the variables used in the regressions, with a VIF_max_<5, classed as an acceptable level of correlation [13]. The models were applied to changes in phases 1 and 2, relative to baseline periods. Statistical analyses of the key variables identified in the regression were performed using Mixed ANOVAs to investigate differences in PA change within sub-groups of individuals, using s within-subjects variable of time period (Phase 1, Phase 2).

In addition to understanding change in levels of PA, we also investigated how PA routines had changed. This was achieved through a Sankey network of the main types of PA (e.g., running, gym, outdoor sports) respondents participated in across the three periods analysed. We subsequently, analysed the intention to stick with new (or old) routines post-lockdown, for each sport type.

## RESULTS

A table of demographic data for the full sample (N=1656) and the sub-sample who provided step-count data (N=487) is provided in the Supplementary Information A.

### Perceived change in PA

Perceived change in PA during lockdown Phase 1 was, on average, slightly negative (M=-.30, sd=2.67). However, the distribution of responses was spread widely, highlighting an almost equal split between those who reported a reduction in PA levels (46.0%) and an increase in PA levels (39.9%), with 14.1% reporting no change. For lockdown Phase 2, there was a slight shift towards more people reporting an increase (43.7%) or no change (18.3%) in PA levels compared to the pre-lockdown periods, with 38.0% reporting a decrease (overall M=.09, sd=2.55).

### Change in step count

Prior to the lockdown periods, mean daily step count across the sample (N=487) was 6680.53 ±3310.24. Both lockdown phases had a significant negative impact on mean daily step count (F(1.61, 781.94)=72.838, p<.001), with the mean daily number of steps reducing to a mean of 5157.07 ±3474.58 in Phase 1, and 6197.62 ±4028.07 in Phase 2.

### Factors associated with perceived physical activity change

Factors associated with the perceived change in PA for both Phase 1 and Phase 2 lockdowns, relative to the Baseline period are shown in Figure 1 (VIF_max_=2.8; Phase 1: N=1656, R^2^=0.12; Phase 2: N=1656, R^2^=0.13). The full table of results for all independent variables is provided in Supplementary Information B.

**Figure 1.**
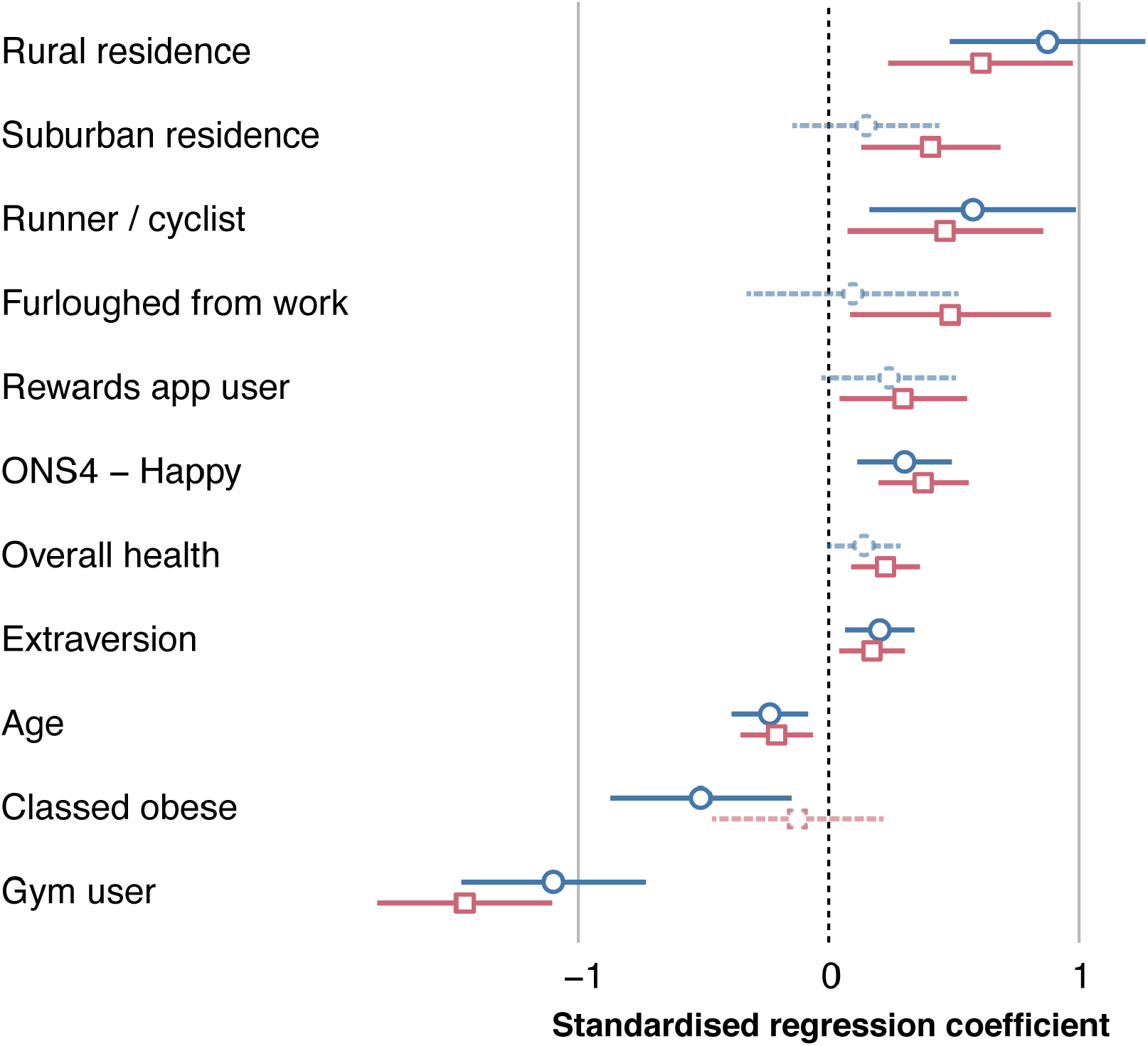
Plot of regression coefficients for the multiple regression model of self-reported perceived change in physical activity for both Phase 1 (blue circle marker) and Phase 2 (pink square marker) lockdown periods, relative to the Baseline period. Variables are only displayed where there is statistical significance in at least one of the phases. Where the variable is only significant in one phase, the non-significant phase is displayed as a dashed line. Error bars represent 95% confidence intervals.

Based on the regression results, we further investigated specific changes relating to the key significant factors identified: weight classification, home location and main type of PA prior to the lockdown.

We further noted the linear positive relationship between perceived change in PA and the ‘happiness’ rating from the Office of National Statistics wellbeing scale [8], and a negative relationship with age, in both phases.

For Weight classification, there was an interaction between Time (Phase 1, Phase 2) and the Weight classification (F(3,1652) = 2.63, p=.049; Figure 2) with people classified as Obese (based on BMI) reporting a significantly larger reduction in perceived change in PA during Phase 1 to those classed as Healthy Weight (p<.001) and Overweight (p=.006). There were no significant differences between weight classifications for Phase 2.

**Figure 2.**
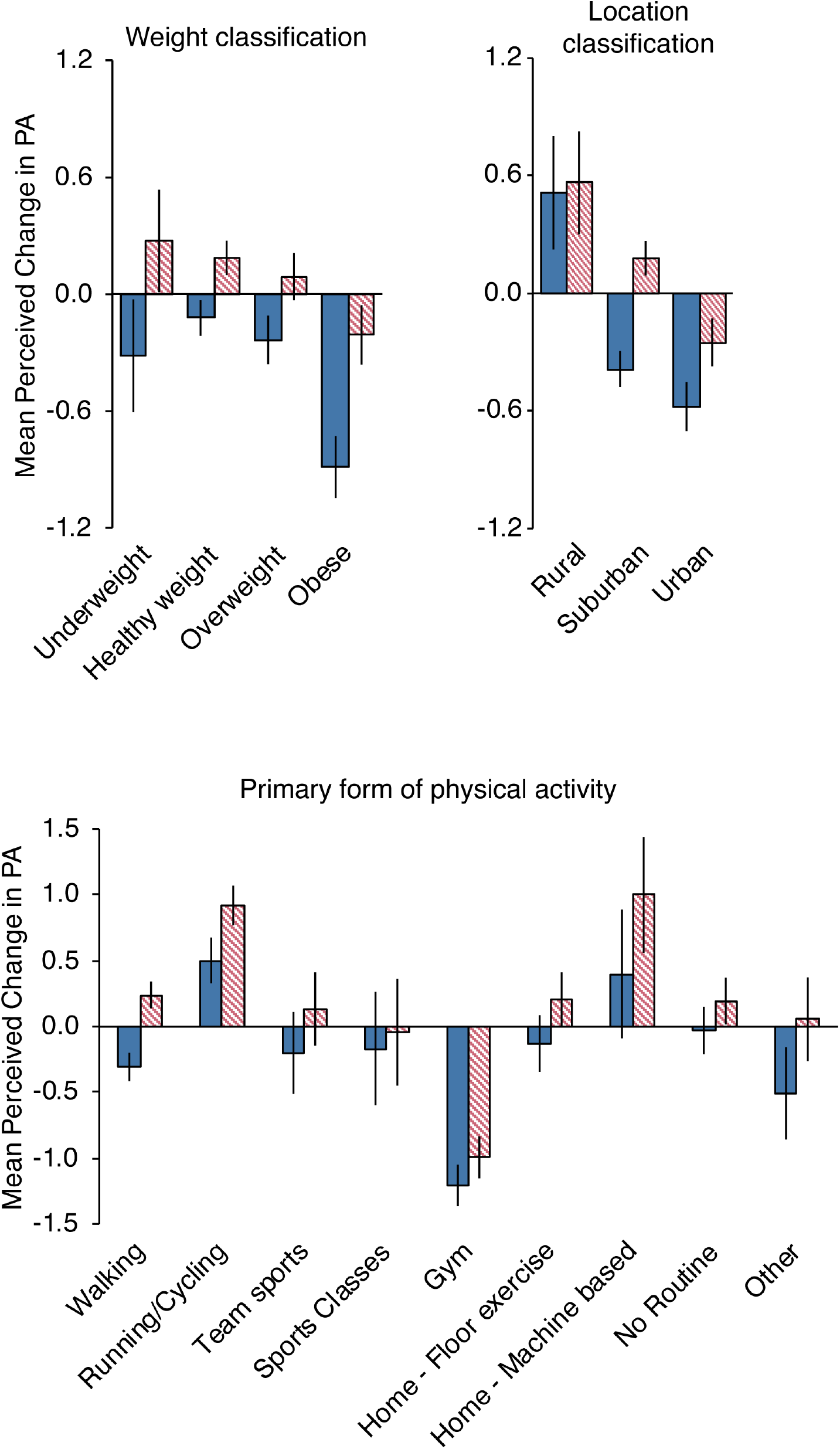
Perceived change in physical activity, by weight classification (top-left), residential location (top-right) and primary PA type prior to restrictions (bottom). Based on a Likert-scale between +5 (substantial increase) and −5 (substantial decrease). Solid blue bars represent Phase 1 period, pink-hatched bars represent Phase 2 period. Error bars represent standard error of the mean.

There was a significant interaction between Time and Residential Location (F(2,1653) = 6.55, p=.001), with those in rural areas reporting increased PA during Phase 1, compared to Suburban (p<.001) and Urban (p<.001) locations, where participants report a reduction in PA (Figure 2). During Phase 2, both those in rural and suburban locations report an increase in PA, with no significant difference between these groups (p=.082). In contrast, those in urban locations continued to report a reduction in PA, which was significantly lower than those in rural (p<.001) and suburban locations (p=.006).

Finally, we found significant differences in perceived PA change based on the main activity type reported by participants prior to the lockdown (F(8,1647)=10.57, p<.001; Figure 2). During Phase 1, while there is little reported change for most activity types, those whose main activity involved the gym prior to the lockdown period, reported a large reduction in PA, which was significantly reduced in comparison to those who did walking (p<001), running/cycling (p<.001), floor exercises (p=.003) or had no specific exercise routine (p<.001). In contrast, those whose main activity was running or cycling prior to the lockdown period, reported a perceived increase in activity during Phase 1, which was significantly higher than those whose main activity was walking (p=.005). Similarly, in Phase 2, those who were primarily gym users prior to the lockdown period continued to report significantly reduced PA compared to all other activity types (p<.05), except for those who did classes or other, unlisted types of activity.

### Factors associated with step-count change

A further multiple regression model was run on the subset of participants who had provided step-count data. We used the log-percentage change in step count for both Phase 1 and Phase 2 lockdown periods, relative to the Baseline period (Table 1) as the dependent variable. The significant factors associated with change in step count (VIF_max_=2.9; Phase 1: N=487, R^2^=0.27; Phase 2: N=487, R^2^=0.25) are shown in Figure 3. For full table of results see Supplementary information C.

**Figure 3.**
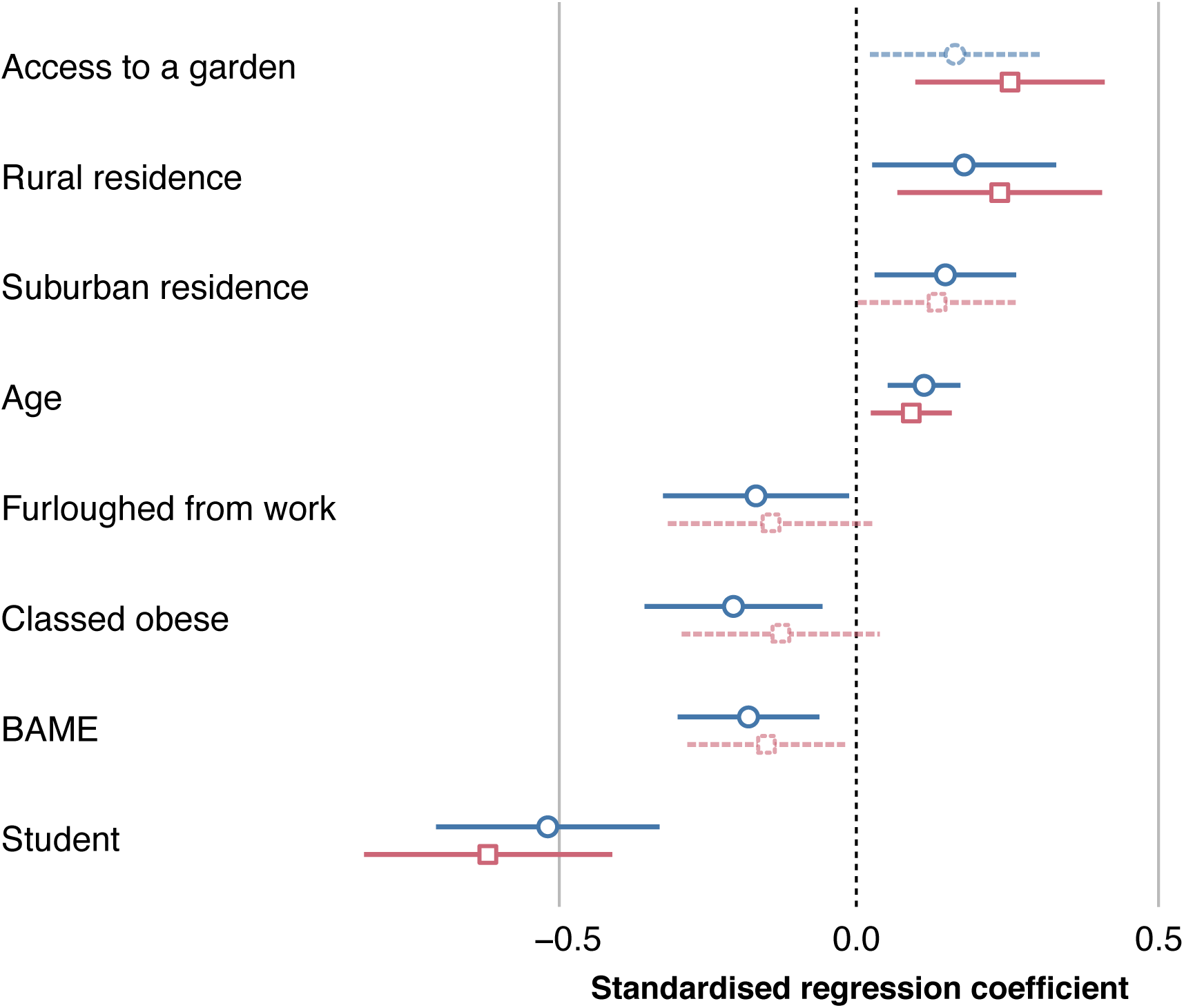
Plot of regression coefficients for the multiple regression model of log-percentage change in mean daily step count for both Phase 1 (blue circle marker) and Phase 2 (pink square marker) lockdown periods, relative to the Baseline period. Variables are only displayed where there is statistical significance in at least one of the phases. Where the variable is only significant in one phase, the non-significant phase is displayed as a dashed line. Error bars represent 95% confidence intervals.

Based on the regression results, we further investigated changes relating to the key factors associated with step count change: weight classification, home location and main type of PA prior to lockdown. We further noted the linear positive relationship between the log-percentage change in step count and age, in both phases.

Although people classed as obese were shown to have a significant negative relationship to change in step count in Phase 1 in the regression (Figure 4), there was no significant main effect of weight classification in the ANOVA analysis (F(3,483)=.235, p=.872).

**Figure 4.**
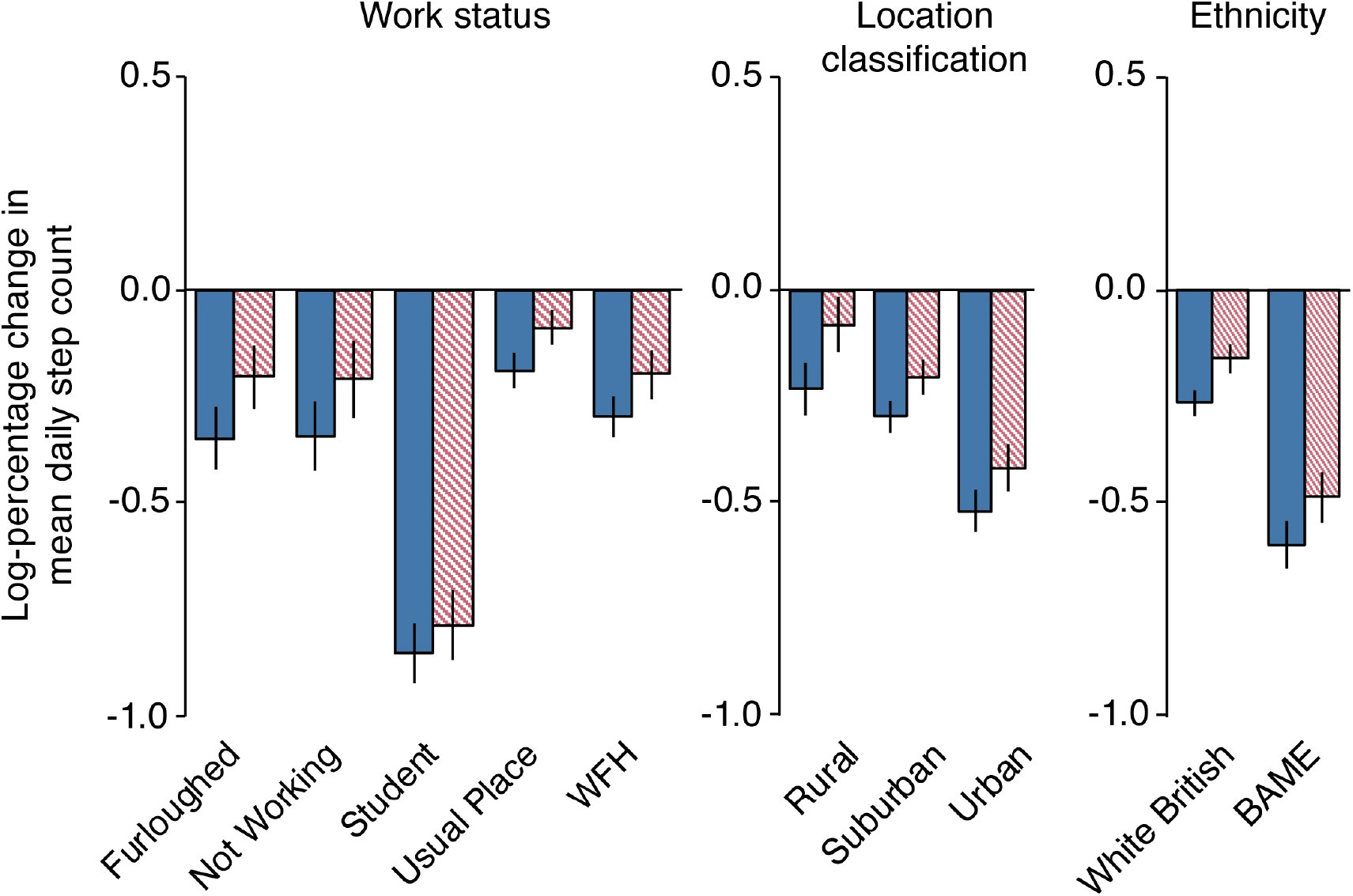
Log-percentage change in mean daily step count for work status (left), residential location (middle) and ethnicity (right). Solid blue bars represent Phase 1 period, pink-hatched bars represent Phase 2 period. Error bars represent standard error of the mean.

As with perceived change, there was a substantial difference in step-count change, dependant on where participants resided (F(2,484)=10.127, p<.001; Figure 4). In particular, those in urban locations had a significantly larger reduction in mean daily step count during both phases than those living in suburban (Phase 1: p<.001; Phase 2: p=.004) or rural locations (Phase 1: p<.001; Phase 2: p<.001). There was no difference between those living in suburban and rural locations for either Phase 1 or 2.

Individuals whose ethnicity was classed as Black, Asian or other minority ethnicity (BAME) showed a significantly larger reduction in mean daily step count during both phases compared to White British (F(1,485)=32.344, p<.001; Figure 4).

We finally investigated the differences in mean daily step count based on work status (F(4,482)=18.42, p<.001; Figure 4). During both phases we observed a substantial reduction in step count in those classified as students, which was significantly lower than all other groups (p<.05). In addition, those who were furloughed at the time of the survey, had a higher reduction in mean daily step count, than those who were working at their usual place of work (other than working from home; p=.046).

### Change in routines

The main exercise routine of 25.9% of the sample was subsequently restricted due to the lockdown rules put in place after 23^rd^ March 2020. Of the remainder, 62.5% took part in activities that weren’t subsequently restricted and 11.6% had no routine prior to the restrictions. The proportion of those participating in unrestricted activities increasing to 83.3% and 85.1% in Phases 1 and 2, respectively. However, there was also a small increase in those reporting no specific PA routine during Phase 1 (14.1%) and Phase 2 (12.5%). The changes in routine are further visualised in the Sankey diagram (Figure 5).

**Figure 5.**
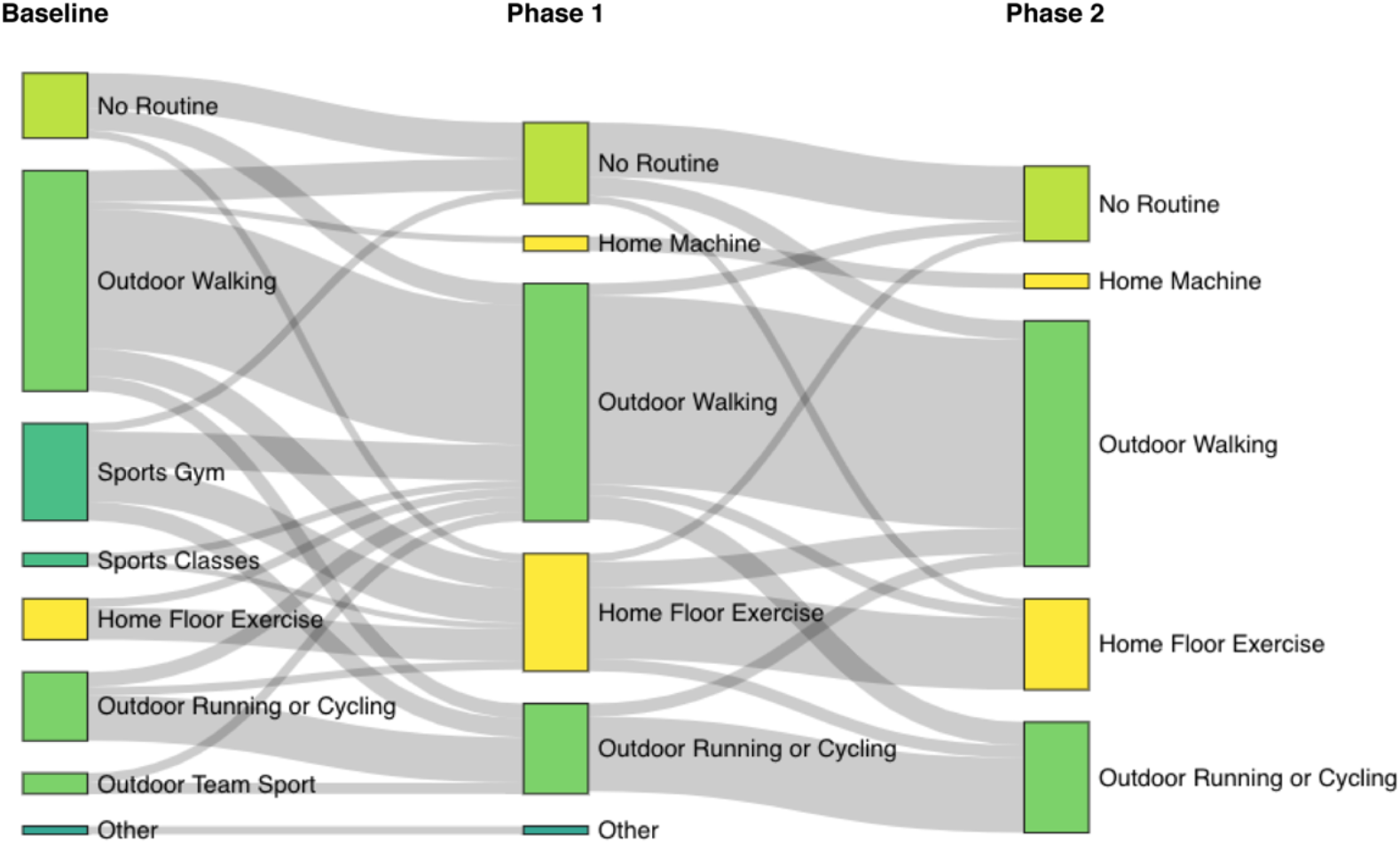
Sankey diagram showing the switch of main physical activity type across the lockdown periods. Block sizes represent proportion of the sample undertaking the activity type. To increase clarity, counts of <15 are not displayed on the diagram.

Finally, we asked those who had changed to a new routine, due to their previous primary activity being restricted, whether they planned to stick to it (Table 2).

**Table 2.** Proportion of respondents who stated they were likely to stick with their new routine once restrictions were lifted or return to their old routine. The results are shown only for respondents who, prior to the lockdown periods, were participating in activities that were subsequently restricted, or had no routine.

| Activity Type | Stick to new routine (percentage) | Stick to old routine (percentage) | Unsure (percentage) |
| --- | --- | --- | --- |
| Gym | 31.2 | 63.5 | 5.3 |
| Fitness classes | 28.6 | 66.7 | 4.8 |
| Outdoor team sports | 43.0 | 44.3 | 12.7 |
| No Routine | 58.3 | 16.7 | 25.0 |

## DISCUSSION

Overall, we found that average step count, measured objectively from smartphone data, reduced during both phases of lockdown in comparison to the period in February prior to the lockdown periods. Taking into account seasonality, this reduction is even more substantial as typically step-count would rise through the months of March-May, when the weather becomes more favourable [14,15]. Similar results have been reported internationally from other app-based measures of step-count recently [16], corroborating the impact lockdown had on activity levels. Here we have provided a more detailed insight using a comprehensive questionnaire in parallel with the step-count data from a large sample to understand which groups have shown the greatest reductions.

While step-count provides a useful objective indicator of PA levels, it must be recognised this only captures a single modality of activity; therefore, we further captured perceived change in PA levels from respondents. This subjective data also provided a larger sample for analysis. Importantly, the distribution of responses differed to that resulting from step-count analyses, with a mean value close to zero in both periods of lockdown. This highlighted a clear split, between those who considered their levels of PA had increased during the lockdown periods, and those who considered it had decreased. The subsequent analyses have highlighted the stark contrasts within groups defined by the demographic, lifestyle and health factors associated with increases or decreases in PA during the UK lockdown periods.

### Residential environment

One of the factors that was significant for both subjective and objective measures of PA was the residential location of participants; those living in rural and suburban locations showed a perceived increase in PA and a lower reduction in step count in at least one lockdown phase. In contrast, urban residents reported a reduction in perceived PA as well as step count in both phases. The restriction to all but essential travel and closure of sports/gym facilities resulted in highly localised PA options [17]. This has emphasised inequalities between rural locations with open green space and urban environments with limited green space and poor walking infrastructure [18], that cannot support localised PA [17].

### Health factors

A concerning finding was that those classed as obese, and hence already likely to have sedentary lifestyles, were reporting substantially lower levels of exercise than those in other weight groups. While there was no significant difference to other weight groups in percentage step-count reduction, the obese group also reported an overall reduction in step-count during both phases. It has been identified that those classed as obese are at higher risk of developing complications from COVID-19 [19,20], with the impact of reduced PA on the immune system being a contributing factor [20].

Hence, it is concerning that lockdown restrictions could potentially exacerbate this group’s vulnerability due to further reductions in PA in an already inactive group. Another group, evidenced to be at higher risk from COVID-19 are those from BAME populations [21,22]. Again, we found a stark contrast in PA levels, in terms of reduced step-count, in those from BAME groups compared to those identifying as White British.

In addition to physical health, PA has a positive impact on mental health, with those participating in higher levels of activity having lower odds of developing depressive symptoms [23]. It is interesting therefore, that there was a significant positive correlation between perceived change in PA and the happiness rating from the ONS4 scale. This highlights, and further corroborates similar studies (e.g.,

[24] that, on average, those who had increased PA during lockdown were also more likely to be happier during that period. In contrast, a sudden stop in PA in previously active people risks increasing depressive symptoms within a short period of time [25]. Hence, the sudden reduction in activity levels, from those who have been unable to maintain their usual routine may have exacerbated this relationship between mood and change in activity, which has been shown to have deteriorated nationally in the UK during the lockdown [26].

### Age

We observed an interesting contrast of age within the analyses, with a negative correlation of age with perceived change in PA, in line with other research [27], versus a positive correlation of age with step-count. From this we can infer that older age groups feel that their overall PA levels have reduced more in comparison to younger groups during the lockdown periods. However, older groups were possibly more likely to switch to walking or running activities resulting in a smaller reduction in step count than younger groups, also mirrored by the significant reduction in students’ step-count compared to other work groups. Given the sample demographic however, it is important to contextualise these results by highlighting that older groups here are more likely to be defined as middle-aged adults, as opposed to older adults per se (<6% of the sample were aged over 55 years).

### Exercise types

It is interesting how the primary form of PA (prior to restrictions) impacted perceived change in PA during the lockdown periods. Those who were primarily runners/cyclists tended to report increased levels of activity during the restrictions, possibly having more opportunities to undertake this opportunity. Gym users reported by far the biggest reduction in perceived PA during the restrictions. This highlights the reliance and habituation gym users have on these facilities, which were closed during the lockdown periods. It is clear that, whilst most switched to new outdoor or home-based activities during the closures, they did not feel they were achieving the same level of exercise as their previous routines. This is further reflected in the fact that two-thirds of gym users planned to return to their previous PA routines, once restrictions were lifted. A similar proportion planned to return to fitness classes, and highlights the strong reliance and affiliation to these types of PA. The lockdown period did provide some opportunity to those who previously reported having no specific PA routine. Of those who developed a routine during the restrictions, over half planned to continue with this new routine once restrictions had lifted.

### Limitations

The sample of respondents lacked older adults – less than 6% of the full sample were over the age of 55 years. Therefore, we cannot generalise our results to older age groups. However, the large sample we collected did allow us to provide a comprehensive insight into how the pandemic related restrictions have impacted PA across different demographic groups.

## CONCLUSION

The results from the study highlight the dichotomy the impact has had on PA routines. Crucially, groups at high risk of complications from COVID-19 appear to be also impacted in terms of substantial reduction in PA. More specifically, those who are obese are at risk of further reducing already low activity levels; if the impact of continued restrictions has a long-term effect on routines, this further reduction could become habitualised. Therefore, we suggest that interventions are required to support these groups, to ensure they have access and motivation to participate in physical activities, whether this is home based or outdoors.

On the other hand, we have seen some groups develop new routines and increase (perceived) levels of PA during the restrictions. Support should also be provided to these groups to maintain these new routines to ensure they are long lasting, and hence be beneficial to both their mental and physical health.

## Data Availability

Anonymised questionnaire responses and step count data are available from the OSF respository: https://osf.io/b4wz8.

## ACKNOWLEDGEMENTS

We thank Shaun Azam and the team at Sweatco Ltd, for their assistance with data collection through the Sweatcoin app. We further thank Dr Lukasz Walasek for his assistance with the design and ethical approval applications for this project.

## AUTHOR CONTRIBUTIONS

MTE, CJ, IV designed the study and collected the data; MTE, VE, MS analysed the data; All authors contributed to writing of the manuscript.

## References

1 Prime Minister’s statement on coronavirus (COVID-19): 23 March 2020. GOV.UK. https://www.gov.uk/government/speeches/pm-address-to-the-nation-on-coronavirus-23-march-2020 (accessed 15 Dec 2020).

2 Prime Minister’s statement on coronavirus (COVID-19): 10 May 2020. GOV.UK. https://www.gov.uk/government/speeches/pm-address-to-the-nation-on-coronavirus-10-may-2020 (accessed 15 Dec 2020).

3 Woods JA, Hutchinson NT, Powers SK, et al. The COVID-19 pandemic and physical activity. Sports Medicine and Health Science 2020;2:55–64. doi:10.1016/j.smhs.2020.05.006

4 Coronavirus and homeworking in the UK - Office for National Statistics. https://www.ons.gov.uk/employmentandlabourmarket/peopleinwork/employmentandemployeetypes/bulletins/coronavirusandhomeworkingintheuk/april2020 (accessed 15 Dec 2020).

5 Comparison of furloughed jobs data - Office for National Statistics. https://www.ons.gov.uk/businessindustryandtrade/business/businessservices/articles/comparisonoffurloughedjobsdata/maytojuly2020 (accessed 15 Dec 2020).

6 Derlyatka A, Fomenko O, Eck F, et al. Bright spots, physical activity investments that work: Sweatcoin: a steps generated virtual currency for sustained physical activity behaviour change. Br J Sports Med 2019;:bjsports-2018–099739. doi:10.1136/bjsports-2018-099739

7 Palan S, Schitter C. Prolific.ac—A subject pool for online experiments. Journal of Behavioral and Experimental Finance 2018;17:22–7. doi:10.1016/j.jbef.2017.12.004

8 Waldron S. Measuring subjective wellbeing in the UK. Office for National Statistics 2010. http://www.mas.org.uk/uploads/artlib/measuring-subjective-wellbeing-in-the-uk.pdf (accessed 15 Dec 2020).

9 De Raad B. The Big Five Personality Factors: The psycholexical approach to personality. Ashland, OH, US: Hogrefe & Huber Publishers 2000.

10 Gosling SD, Rentfrow PJ, Swann WB. A very brief measure of the Big-Five personality domains. Journal of Research in Personality 2003;37:504–28. doi:10.1016/S0092-6566(03)00046-1

11 General practice physical activity questionnaire (GPPAQ). GOV.UK. 2013.https://www.gov.uk/government/publications/general-practice-physical-activity-questionnaire-gppaq (Archived by WebCite® at http://www.webcitation.org/731Pr3rrM) (accessed 19 Jun 2018).

12 R Core Team. R: A Language and Environment for Statistical Computing. Vienna, Austria: R Foundation for Statistical Computing 2019. https://www.R-project.org/

13 Daoud JI. Multicollinearity and Regression Analysis. J Phys: Conf Ser 2017;949:012009. doi:10.1088/1742-6596/949/1/012009

14 Tucker P, Gilliland J. The effect of season and weather on physical activity: A systematic review. Public Health 2007;121:909–22. doi:10.1016/j.puhe.2007.04.009

15 Tudor-Locke C, Bassett DR, Swartz AM, et al. A Preliminary study of one year of pedometer self-monitoring. ann behav med 2004;28:158–62. doi:10.1207/s15324796abm2803_3

16 Tison GH, Avram R, Kuhar P, et al. Worldwide Effect of COVID-19 on Physical Activity: A Descriptive Study. Ann Intern Med 2020;173:767–70. doi:10.7326/M20-2665

17 McDougall CW, Brown C, Thomson C, et al. From one pandemic to another: emerging lessons from COVID-19 for tackling physical inactivity in cities. Cities & Health 2020;0:1–4. doi:10.1080/23748834.2020.1785165

18 McCormack G, Giles-Corti B, Lange A, et al. An update of recent evidence of the relationship between objective and self-report measures of the physical environment and physical activity behaviours. Journal of Science and Medicine in Sport 2004;7:81–92. doi:10.1016/S1440-2440(04)80282-2

19 Lighter J, Phillips M, Hochman S, et al. Obesity in Patients Younger Than 60 Years Is a Risk Factor for COVID-19 Hospital Admission. Clinical Infectious Diseases 2020;71:896–7. doi:10.1093/cid/ciaa415

20 Kimura T, Namkoong H. Susceptibility of the obese population to COVID-19. International Journal of Infectious Diseases 2020;101:380–1. doi:10.1016/j.ijid.2020.10.015

21 Bhatia M. COVID-19 and BAME Group in the United Kingdom. The International Journal of Community and Social Development 2020;2:269–72. doi:10.1177/2516602620937878

22 Niedzwiedz CL, O’Donnell CA, Jani BD, et al. Ethnic and socioeconomic differences in SARS-CoV-2 infection: prospective cohort study using UK Biobank. medRxiv 2020;:2020.04.22.20075663. doi:10.1101/2020.04.22.20075663

23 Schuch FB, Vancampfort D, Firth J, et al. Physical Activity and Incident Depression: A Meta-Analysis of Prospective Cohort Studies. AJP 2018;175:631–48. doi:10.1176/appi.ajp.2018.17111194

24 Jacob L, Tully MA, Barnett Y, et al. The relationship between physical activity and mental health in a sample of the UK public: A cross-sectional study during the implementation of COVID-19 social distancing measures. Mental Health and Physical Activity 2020;19:100345. doi:10.1016/j.mhpa.2020.100345

25 Edwards MK, Loprinzi PD. Effects of a Sedentary Behavior-Inducing Randomized Controlled Intervention on Depression and Mood Profile in Active Young Adults. Mayo Clin Proc 2016;91:984–98. doi:10.1016/j.mayocp.2016.03.021

26 Pierce M, Hope H, Ford T, et al. Mental health before and during the COVID-19 pandemic: a longitudinal probability sample survey of the UK population. The Lancet Psychiatry 2020;7:883– 92. doi:10.1016/S2215-0366(20)30308-4

27 Rogers NT, Waterlow NR, Brindle H, et al. Behavioral Change Towards Reduced Intensity Physical Activity Is Disproportionately Prevalent Among Adults With Serious Health Issues or Self-Perception of High Risk During the UK COVID-19 Lockdown. Front Public Health 2020;8. doi:10.3389/fpubh.2020.575091

