## Supplementary Info B for "Stay Home and Stay Active? The impact of stay-at-home restrictions on physical activity routines in the UK during the COVID-19 pandemic" (from another version of this preprint: version 1, 2021-02-03)

**Supplementary Information – Perceived Change in Physical Activity Regression Results**

Victoria Eshelby, Muhammed Sogut, Catherine Jolly, Ivo Vlaev, Mark T. Elliott

**Dependent Variable:** Perceived change in physical activity – Phase 1.
**Sample**: N=1656
**Model Fit**: $R^2 = 0.12$.

| Variable | Est. | S.E. | t-statistic | p-value (significant in bold) |
|---|---|---|---|---|
| isSWCSample | 0.24 | 0.14 | 1.70 | 0.09 |
| **age** | **-0.24** | **0.08** | **-3.02** | **0.00** |
| isSuburb | 0.15 | 0.15 | 0.96 | 0.34 |
| **isRural** | **0.87** | **0.19** | **4.49** | **0.00** |
| hasGarden | 0.12 | 0.18 | 0.68 | 0.50 |
| hasChildren | -0.22 | 0.14 | -1.62 | 0.11 |
| isUnderweight | -0.10 | 0.29 | -0.36 | 0.72 |
| isOverweight | -0.02 | 0.16 | -0.11 | 0.91 |
| **isObese** | **-0.51** | **0.19** | **-2.66** | **0.01** |
| isBAME | -0.27 | 0.17 | -1.58 | 0.12 |
| ons4.lifesat | 0.15 | 0.11 | 1.38 | 0.17 |
| ons4.worthwhile | -0.01 | 0.11 | -0.10 | 0.92 |
| **ons4.happy** | **0.30** | **0.10** | **2.96** | **0.00** |
| ons4.anxiety | 0.04 | 0.08 | 0.56 | 0.58 |
| health | 0.14 | 0.08 | 1.82 | 0.07 |
| **.data_Outdoor.Running.or.Cycling** | **0.57** | **0.21** | **2.74** | **0.01** |
| .data_Outdoor.Team.Sport | -0.37 | 0.33 | -1.13 | 0.26 |
| .data_Outdoor.Other | -0.45 | 0.62 | -0.73 | 0.46 |
| **.data_Sports.Gym** | **-1.10** | **0.20** | **-5.51** | **0.00** |
| .data_Sports.classes | -0.03 | 0.42 | -0.06 | 0.95 |
| .data_Home.floor | 0.23 | 0.23 | 1.00 | 0.32 |
| .data_Home.machine | 0.37 | 0.50 | 0.75 | 0.46 |
| .data_No.routine | 0.35 | 0.21 | 1.68 | 0.09 |
| .data_Other | -0.30 | 0.43 | -0.71 | 0.48 |
| work.isFurloughed | 0.09 | 0.22 | 0.43 | 0.67 |
| work.isWFH | -0.15 | 0.19 | -0.81 | 0.42 |
| work.notWorkRet | -0.22 | 0.22 | -1.02 | 0.31 |
| work.student | -0.36 | 0.25 | -1.48 | 0.14 |
| active.commuterTRUE | -0.16 | 0.15 | -1.05 | 0.29 |
| had.covid | 0.10 | 0.22 | 0.46 | 0.65 |
| shielding.adhering | 0.45 | 0.33 | 1.34 | 0.18 |
| shielding.notadh | 0.65 | 0.48 | 1.36 | 0.17 |

| | | | | |
|---|---|---|---|---|
| distancing.comply.num | 0.06 | 0.07 | 0.94 | 0.35 |
| covid.anxiety.num | -0.03 | 0.06 | -0.48 | 0.63 |
| discount.long | 0.05 | 0.14 | 0.33 | 0.74 |
| **extraversion** | **0.20** | **0.07** | **2.82** | **0.00** |
| agreeable | -0.12 | 0.07 | -1.80 | 0.07 |
| conscient | 0.07 | 0.07 | 1.00 | 0.32 |
| emostab | -0.05 | 0.08 | -0.56 | 0.58 |
| openness | -0.13 | 0.07 | -1.85 | 0.06 |

**Dependent Variable:** Perceived change in physical activity – Phase 2.
**Sample**: N=1656
**Model Fit**: $R^2 = 0.13$.

| Variable | Est. | S.E. | t-statistic | p-value (significant in bold) |
|---|---|---|---|---|
| **isSWCSample** | **0.30** | **0.13** | **2.21** | **0.03** |
| **age** | **-0.21** | **0.07** | **-2.84** | **0.00** |
| **isSuburb** | **0.41** | **0.15** | **2.75** | **0.01** |
| **isRural** | **0.61** | **0.19** | **3.18** | **0.00** |
| hasGarden | 0.01 | 0.17 | 0.07 | 0.95 |
| hasChildren | -0.05 | 0.13 | -0.40 | 0.69 |
| isUnderweight | 0.23 | 0.26 | 0.89 | 0.38 |
| isOverweight | 0.02 | 0.15 | 0.17 | 0.87 |
| isObese | -0.13 | 0.18 | -0.70 | 0.48 |
| isBAME | -0.07 | 0.16 | -0.45 | 0.65 |
| ons4.lifesat | 0.04 | 0.11 | 0.38 | 0.70 |
| ons4.worthwhile | 0.01 | 0.11 | 0.12 | 0.91 |
| **ons4.happy** | **0.38** | **0.10** | **3.82** | **0.00** |
| ons4.anxiety | 0.02 | 0.08 | 0.21 | 0.83 |
| **health** | **0.23** | **0.07** | **3.08** | **0.00** |
| **.data_Outdoor.Running.or.Cycling** | **0.46** | **0.18** | **2.51** | **0.01** |
| .data_Outdoor.Team.Sport | -0.50 | 0.32 | -1.56 | 0.12 |
| .data_Outdoor.Other | 0.05 | 0.55 | 0.09 | 0.93 |
| **.data_Sports.Gym** | **-1.45** | **0.19** | **-7.67** | **0.00** |
| .data_Sports.classes | -0.45 | 0.42 | -1.07 | 0.29 |
| .data_Home.floor | 0.01 | 0.23 | 0.06 | 0.95 |
| .data_Home.machine | 0.48 | 0.43 | 1.10 | 0.27 |
| .data_No.routine | -0.01 | 0.20 | -0.07 | 0.94 |
| .data_Other | -0.51 | 0.39 | -1.31 | 0.19 |
| **work.isFurloughed** | **0.48** | **0.21** | **2.34** | **0.02** |
| work.isWFH | -0.04 | 0.18 | -0.21 | 0.83 |
| work.notWorkRet | -0.03 | 0.21 | -0.15 | 0.88 |
| work.student | -0.29 | 0.24 | -1.20 | 0.23 |

| | | | | |
|---|---|---|---|---|
| active.commuterTRUE | -0.21 | 0.14 | -1.50 | 0.13 |
| had.covid | 0.05 | 0.19 | 0.25 | 0.80 |
| shielding.adhering | 0.16 | 0.34 | 0.46 | 0.65 |
| shielding.notadh | 0.34 | 0.47 | 0.73 | 0.46 |
| distancing.comply.num | 0.11 | 0.06 | 1.72 | 0.09 |
| covid.anxiety.num | 0.02 | 0.06 | 0.38 | 0.71 |
| discount.long | 0.08 | 0.13 | 0.63 | 0.53 |
| **extraversion** | **0.17** | **0.07** | **2.45** | **0.01** |
| agreeable | -0.12 | 0.06 | -1.81 | 0.07 |
| conscient | 0.06 | 0.07 | 0.87 | 0.39 |
| emostab | -0.05 | 0.08 | -0.61 | 0.54 |
| openness | -0.05 | 0.07 | -0.76 | 0.45 |
