## Supplementary Info C for "Stay Home and Stay Active? The impact of stay-at-home restrictions on physical activity routines in the UK during the COVID-19 pandemic" (from another version of this preprint: version 1, 2021-02-03)

**Supplementary Information – Step Count Change Regression Results**

Victoria Eshelby, Muhammed Sogut, Catherine Jolly, Ivo Vlaev, Mark T. Elliott

**Dependent Variable:** Step-count change in physical activity – Phase 1.
**Sample**: N=487
**Model Fit**: $R^2 = 0.27$.

| Variable | Est. | S.E. | t-statistic | p-value (significant in bold) |
|---|---|---|---|---|
| **age** | **0.11** | **0.03** | **3.70** | **0.00** |
| **isSuburb** | **0.15** | **0.06** | **2.43** | **0.02** |
| **isRural** | **0.18** | **0.08** | **2.29** | **0.02** |
| hasGarden | 0.17 | 0.09 | 1.85 | 0.07 |
| hasChildren | -0.02 | 0.05 | -0.28 | 0.78 |
| isUnderweight | -0.03 | 0.17 | -0.21 | 0.83 |
| isOverweight | -0.04 | 0.06 | -0.57 | 0.57 |
| **isObese** | **-0.21** | **0.08** | **-2.64** | **0.01** |
| **isBAME** | **-0.18** | **0.07** | **-2.67** | **0.01** |
| ons4.lifesat | -0.01 | 0.04 | -0.28 | 0.78 |
| ons4.worthwhile | 0.00 | 0.05 | 0.00 | 1.00 |
| ons4.happy | -0.02 | 0.04 | -0.54 | 0.59 |
| ons4.anxiety | -0.01 | 0.03 | -0.34 | 0.74 |
| health | 0.00 | 0.03 | 0.01 | 1.00 |
| .data_Outdoor.RunCycling | -0.05 | 0.08 | -0.64 | 0.52 |
| .data_Outdoor.Team | 0.03 | 0.12 | 0.26 | 0.79 |
| .data_Outdoor.Other | -0.07 | 0.15 | -0.45 | 0.66 |
| .data_Sports.Gym | 0.13 | 0.07 | 1.78 | 0.08 |
| .data_Sports.classes | -0.19 | 0.14 | -1.33 | 0.19 |
| .data_Home.floor | -0.18 | 0.12 | -1.50 | 0.13 |
| .data_Home.machine | 0.06 | 0.17 | 0.34 | 0.74 |
| .data_No.routine | -0.01 | 0.10 | -0.09 | 0.93 |
| .data_Other | 0.05 | 0.15 | 0.33 | 0.74 |
| **work.isFurloughed** | **-0.17** | **0.08** | **-2.05** | **0.04** |
| work.isWFH | -0.12 | 0.06 | -1.84 | 0.07 |
| work.notWorkRet | -0.17 | 0.10 | -1.62 | 0.11 |
| **work.student** | **-0.52** | **0.10** | **-5.00** | **0.00** |
| active.commuterTRUE | -0.02 | 0.06 | -0.30 | 0.76 |
| had.covid | 0.00 | 0.08 | -0.02 | 0.98 |
| shielding.adhering | -0.39 | 0.24 | -1.59 | 0.11 |
| shielding.notadh | -0.05 | 0.12 | -0.41 | 0.68 |
| distancing.comply.num | 0.05 | 0.03 | 1.93 | 0.05 |
| covid.anxiety.num | -0.02 | 0.02 | -0.71 | 0.48 |

| | | | | |
|---|---|---|---|---|
| discount.long | -0.04 | 0.06 | -0.65 | 0.52 |
| extraversion | 0.04 | 0.03 | 1.49 | 0.14 |
| agreeable | -0.04 | 0.03 | -1.50 | 0.13 |
| conscient | 0.01 | 0.03 | 0.50 | 0.62 |
| emostab | 0.02 | 0.04 | 0.49 | 0.62 |
| openness | 0.02 | 0.03 | 0.69 | 0.49 |

**Dependent Variable:** Step-count change in physical activity – Phase 2.
**Sample**: N=487
**Model Fit**: $R^2 = 0.25$.

| Variable | Est. | S.E. | t-statistic | p-value (significant in bold) |
|---|---|---|---|---|
| **age** | **0.09** | **0.03** | **2.78** | **0.01** |
| isSuburb | 0.14 | 0.07 | 1.92 | 0.06 |
| **isRural** | **0.24** | **0.09** | **2.67** | **0.01** |
| **hasGarden** | **0.26** | **0.09** | **2.91** | **0.00** |
| hasChildren | -0.02 | 0.06 | -0.37 | 0.71 |
| isUnderweight | 0.22 | 0.20 | 1.13 | 0.26 |
| isOverweight | -0.06 | 0.07 | -0.80 | 0.42 |
| isObese | -0.13 | 0.08 | -1.51 | 0.13 |
| isBAME | -0.15 | 0.08 | -1.97 | 0.05 |
| ons4.lifesat | -0.04 | 0.05 | -0.81 | 0.42 |
| ons4.worthwhile | 0.02 | 0.05 | 0.35 | 0.73 |
| ons4.happy | -0.02 | 0.04 | -0.41 | 0.68 |
| ons4.anxiety | 0.01 | 0.03 | 0.21 | 0.83 |
| health | 0.02 | 0.04 | 0.65 | 0.52 |
| .data_Outdoor.RunCycling | -0.05 | 0.08 | -0.62 | 0.54 |
| .data_Outdoor.Team | 0.05 | 0.12 | 0.39 | 0.70 |
| .data_Outdoor.Other | -0.33 | 0.22 | -1.52 | 0.13 |
| .data_Sports.Gym | 0.11 | 0.08 | 1.29 | 0.20 |
| .data_Sports.classes | -0.10 | 0.22 | -0.47 | 0.64 |
| .data_Home.floor | -0.16 | 0.14 | -1.14 | 0.25 |
| .data_Home.machine | 0.12 | 0.19 | 0.64 | 0.52 |
| .data_No.routine | -0.02 | 0.14 | -0.17 | 0.87 |
| .data_Other | 0.04 | 0.16 | 0.22 | 0.82 |
| work.isFurloughed | -0.14 | 0.09 | -1.60 | 0.11 |
| work.isWFH | -0.11 | 0.07 | -1.54 | 0.12 |
| work.notWorkRet | -0.15 | 0.11 | -1.44 | 0.15 |
| **work.student** | **-0.62** | **0.12** | **-5.19** | **0.00** |
| active.commuterTRUE | -0.04 | 0.07 | -0.56 | 0.58 |

| | | | | |
|---|---|---|---|---|
| had.covid | 0.12 | 0.08 | 1.43 | 0.15 |
| shielding.adhering | -0.22 | 0.17 | -1.33 | 0.18 |
| shielding.notadh | -0.08 | 0.15 | -0.52 | 0.61 |
| distancing.comply.num | 0.03 | 0.03 | 1.11 | 0.27 |
| covid.anxiety.num | 0.01 | 0.03 | 0.28 | 0.78 |
| discount.long | -0.04 | 0.07 | -0.53 | 0.59 |
| extraversion | 0.05 | 0.04 | 1.31 | 0.19 |
| agreeable | -0.03 | 0.03 | -0.87 | 0.38 |
| conscient | 0.03 | 0.03 | 1.06 | 0.29 |
| emostab | 0.00 | 0.04 | 0.11 | 0.91 |
| openness | 0.06 | 0.03 | 1.68 | 0.09 |
